# E-Cigarette Use and Subclinical Cardiac Effects

**DOI:** 10.1101/2020.01.16.20017780

**Authors:** Florian Rader, Mohamad A. Rashid, Trevor Trung Nguyen, Eric Luong, Andy Kim, Elizabeth H. Kim, Robert Elashhoff, Katherine Davoren, Norma B. Moy, Fida Nafeh, C. Noel Bairey Merz, Joseph E. Ebinger, Naomi M. Hamburg, Jonathan R. Lindner, Susan Cheng

## Abstract

**BACKGROUND:** Electronic (e-) cigarettes are marketed as a safer alternative to conventional tobacco cigarettes. Although e-cigarettes contain a lower level of nicotine, the delivery method involves delivering an aerosolized bolus of poorly-characterized ultrafine particles that have unknown cardiovascular effects.

**METHODS:** We studied apparently adult volunteers, free of any chronic disease, including: non-smoking controls, chronic e-cigarette users, and chronic tobacco cigarette smokers. After overnight abstinence, we used myocardial contrast echocardiography to measure acute increases in myocardial blood flow (MBF)induced by ischemic rhythmic handgrip stress, which causes sympathetically-mediated increases in myocardial work and oxygen demand and, in turn, shear stress, nitric oxide production, and coronary endothelial-dependent vasodilation.

**RESULTS:** In non-smoking controls, handgrip stress increased myocardial blood flow, reflecting normal endothelial function. Chronic tobacco cigarette smokers demonstrated stress-induced blunting in myocardial blood flow change, when compared to non-smoking controls. Chronic e-cigarette smokers demonstrated a decrease, rather than increase, in myocardial blood flow change.

**CONCLUSION:** Chronic e-cigarette users demonstrated substantially impaired coronary microvascular endothelial function, even more pronounced than that seen in chronic tobacco cigarette users. These findings suggest that chronic e-cigarette use leads to measurable and persistent adverse vascular effects that are not directly related to nicotine.

## BACKGROUND

E-cigarette (vaping) devices continue to be perceived as a safer alternative to conventional tobacco cigarettes. While providing variable and sometimes lower amounts of nicotine, e-cigarettes rely on a battery-powered aerosolization method that involves delivering with each inhalation a bolus of poorly-characterized small molecules (e.g. ultra-fine particles, heavy metals, volatile organic compounds). Recent reports have now linked e-cigarette use with toxic inhalation syndromes and risk for severe pulmonary disease.^1^ Beyond direct lung injury, inhaled small molecules can rapidly cross the alveolar-capillary barrier and enter into the circulation, potentially causing harm to other end-organs including the heart.

## METHODS

To understand the possible cardiac effects of e-cigarette use, we prospectively studied N=30 apparently healthy adults (mean age 28±4 years, 27% female) who were free of any chronic disease including: self-reported non-smoking controls (n=10), chronic exclusive e-cigarette users (n=10; mean e-cigarette use 3±2 years), and chronic exclusive tobacco cigarette smokers (n=10; mean smoking history 8±2 years). The institutional review board of Cedars-Sinai Medical Center approved all protocols, and each study participant provided written informed consent.

Following overnight abstinence to ensure nicotine has cleared from their system (average half-life=11 hours),^2^ all participants underwent myocardial contrast echocardiography (MCE) perfusion imaging to quantify relative myocardial blood volume (MBVol), microvascular flux rate (β), and blood flow (MBF) according to previously described methods^3^. Because conventional tobacco cigarettes have been firmly established to cause both acute and chronic coronary endothelial dysfunction and thereby initiating coronary atherosclerosis,^4^ MCE was used to directly compare cigarette smoke and e-cigarette vapor exposure on coronary endothelial function.^5^ Echocardiographic assessments were conducted before and after isometric handgrip exercise (**Figure 1**),^6^ a standardized and reliable exercise stress protocol that leads to sympathetically-mediated increases in myocardial work and oxygen demand (MVO_2_) and, in turn, coronary endothelial-dependent vasodilation under normal conditions. Participants who indicated regular use of both cigarettes and e-cigarettes (i.e. “dual users”) were placed into a separate group which also completed the protocol. Their data was not included in this analysis due to redundancy and time constraints.

**Figure 1.**
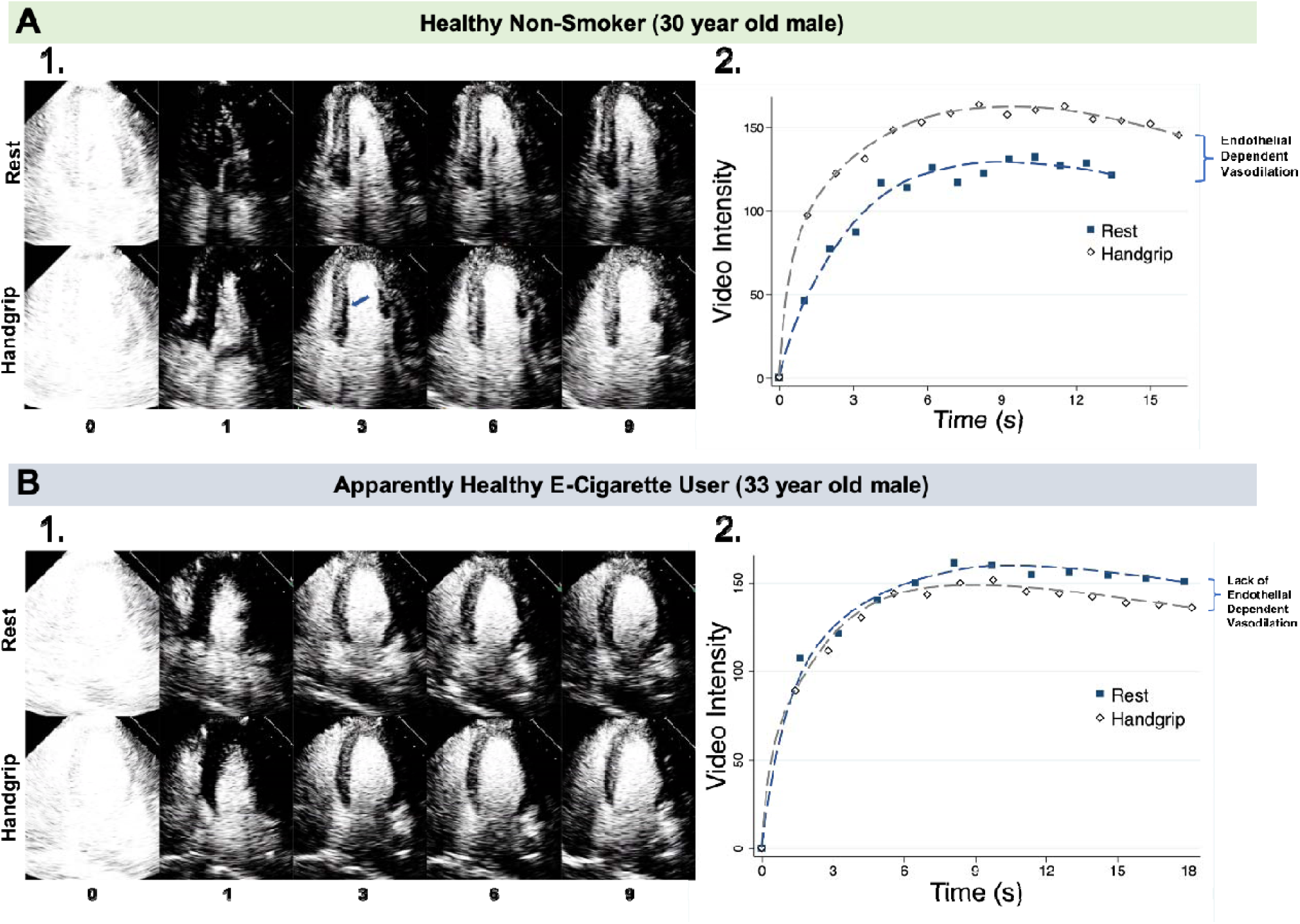
Endothelial-Dependent Coronary Vasodilation Quantified by Myocardial Contrast Echocardiography Before and After Static Handgrip Stress. In **Panel A**, myocardial contrast echocardiography images are shown for a 30 year old healthy non-smoker. After destruction of microbubbles at pulse interval 0, the septal myocardium (arrow) has increased opacification at all post-destructive time intervals during static handgrip at 33% maximal voluntary contraction, indicating increased perfusion. In this participant, both microvascular flux rate and especially the plateau video intensity of the post-destructive time-intensity plot, which represents myocardial blood volume, are clearly increased during static handgrip. Time plots show increase in myocardial blood volume (A) by 21%, microvascular flux rate (β) by 65%, and myocardial blood flow (A x β) by 99%. In **Panel B**, myocardial contrast echocardiography images are shown for a 33 year old apparently healthy e-cigarette user. After destruction of microbubbles at pulse interval 0, we would expect the septal myocardium to opacify similarly to what was seen in the non-smoker; however, it remains dark after microbubble destruction, indicating an impairment of endothelial dependent vasodilation and, thus, perfusion. Time plots show a decrease during static handgrip in myocardial volume by 6%, microvascular flux rate by 25%, and myocardial blood flow by 30%.

## RESULTS

Normally, modest exercise stress induces physiologic increases in MBF through augmentation in myocardial microvascular flux rate with relatively smaller degrees of increase in MBVol. Accordinlgy, in non-smoking controls, exercise produced a statistically significant increase in microvascular flux rate (median: 0.61_pre-test_ vs. 1.14_post-test_ s^-1^) and MBF (median: 84.3_pre-test_ vs. 137.9_post-test_ IU/s) (**Table 1** and **Supplementary Table**). Exercise also increased MBF and flux rate in tobacco users, albeit to a smaller degree than in normal controls; whereas no change in perfusion was seen in e-cigarette users. These results indicate a significant degree of impaired coronary endothelial function that is pronounced in chronic e-cigarette users.

**Table 1.**
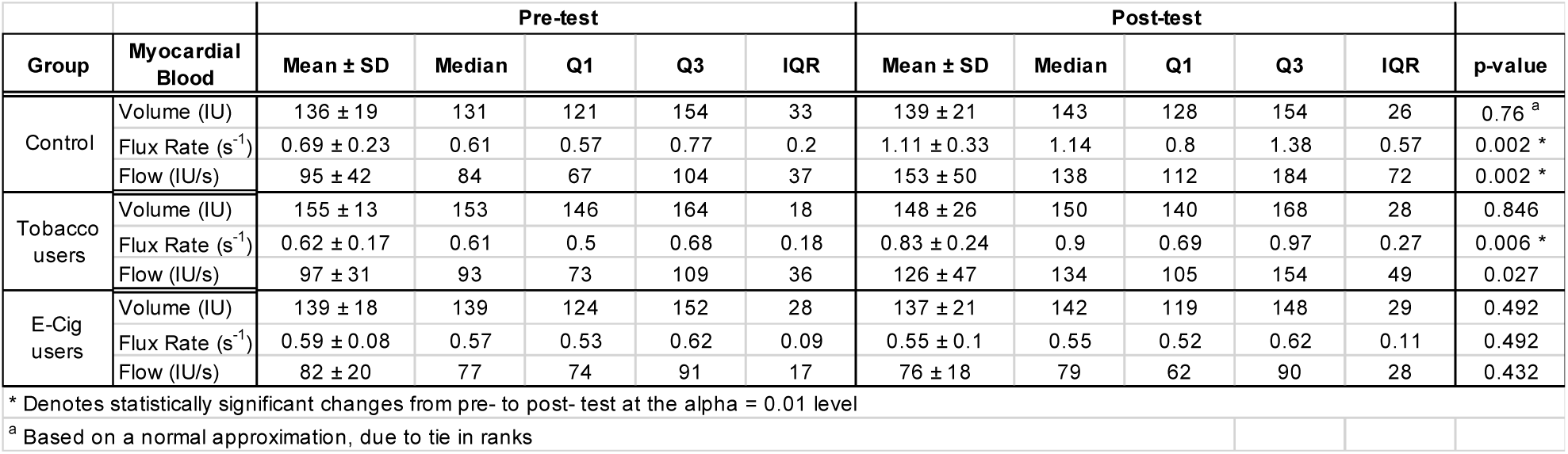
Pre- and Post-Stress Myocardial Blood Flow and Related Measures

To further examine whether changes in myocardial perfusion were different between groups, we compared the percent change in MCE values between groups. The percent changes between the post- and pre-test were compared, as opposed to the raw differences, to account for differences in basal values. Indeed, we detected a significant difference in change of flux rate and MBF between controls and e-cigarette users (**Table 2** and **Figure 2**). In fact, differences were even seen between tobacco users and e-cigarette users.

**Table 2.** Change Between Pre- and Post-Stress Myocardial Blood Flow and Related Measures Across User Groups

| Myocardial Blood | Controls |  |  |  |  | Tobacco users |  |  |  |  | E-cig users |  |  |  |  | p-values |  |  |
| --- | --- | --- | --- | --- | --- | --- | --- | --- | --- | --- | --- | --- | --- | --- | --- | --- | --- | --- |
|  | Mean ± SD | Median | Q1 | Q3 | IQR | Mean ± SD | Median | Q1 | Q3 | IQR | Mean ± SD | Median | Q1 | Q3 | IQR | Control s vs. Tobacc o users | Controls vs. E-cig users | Tobacco vs. E-cig users |
| Volume (IU) | 2.7 ± 12.7 | 0.1 | -2.9 | 11.4 | 14.3 | -4.3 ± 16.8 | -3.9 | -7.2 | 8.3 | 15.5 | -0.9 ± 9.2 | -2.3 | -7 | 3.5 | 10.5 | 0.353 | 0.529 | 0.971 |
| Flux Rate (s) | 66.8 ± 42.8 | 71.5 | 27.3 | 92.2 | 64.9 | 34.7 ± 28.3 | 42.4 | 13.4 | 54.4 | 41.1 | -4.7 ± 20.9 | -0.3 | -24.5 | 5.3 | 29.8 | 0.105 | < 0.001 * | 0.005 * |
| Flow (IU/s) | 72.1 ± 53.2 | 67.8 | 30.5 | 98.4 | 67.9 | 30.5 ± 40.9 | 30.1 | 6.5 | 50.3 | 43.8 | -5.8 ± 20.8 | -4.2 | -20.3 | 5 | 25.3 | 0.089 | < 0.001 * | 0.023 |

\* Denotes a statistically significant difference between in MB measurement between the two groups at an alpha = 0.01 level

**Figure 2.**
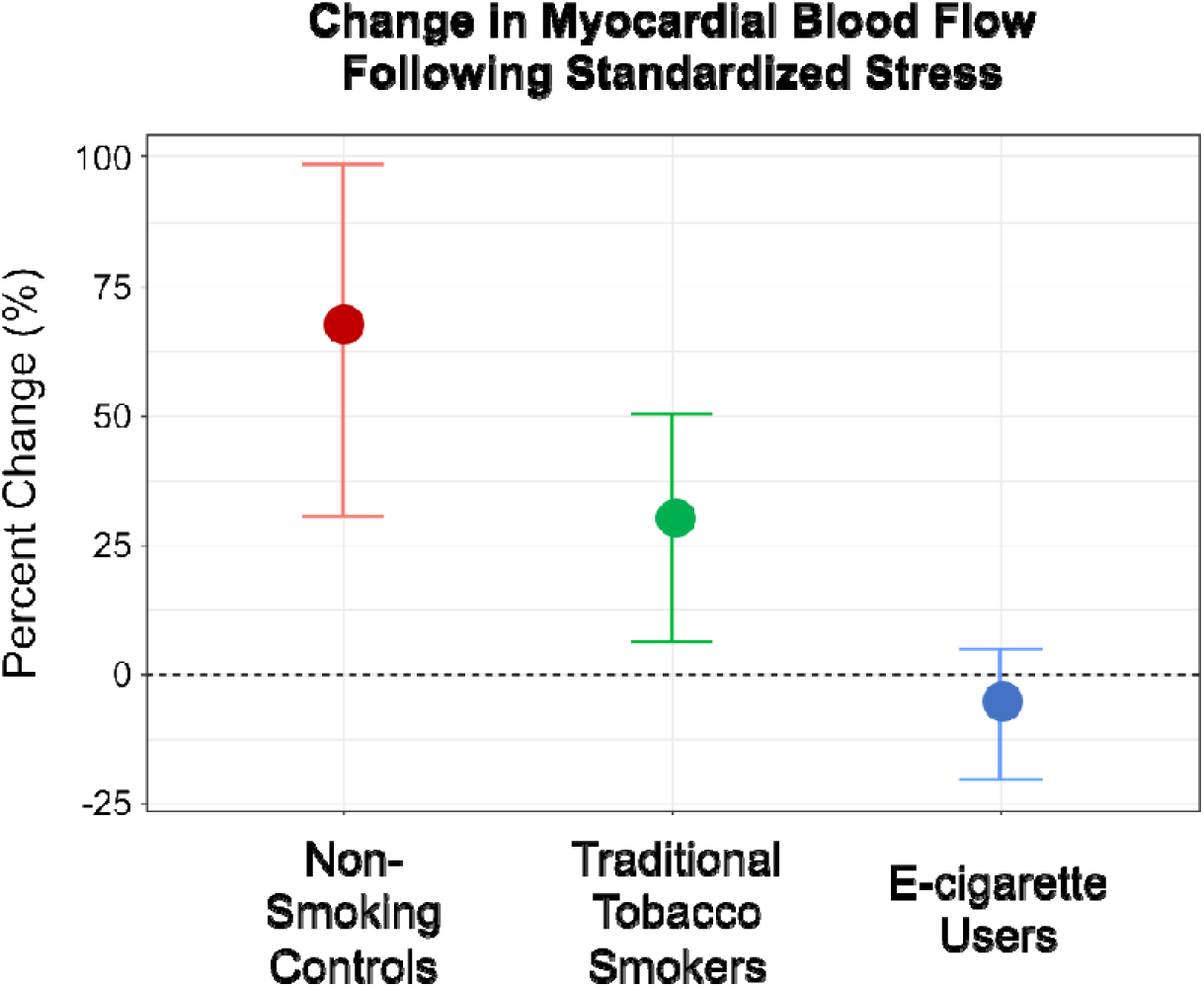
Change in Myocardial Blood Flow Fofllowing Standardized Stress.

## DISCUSSION

Use of conventional combustible cigarettes has long been associated with vascular dysfunction as well as incident cardiovascular disease. In this prospective study, we found evidence of coronary microvascular endothelial dysfunction that was even worse in exclusive e-cigarettes users than in exclusive combustible cigarette users. Importantly, these adverse cardiovascular effects were seen to persist in apparently healthy young adult users. While the longer-term cardiovascular consequences of e-cigarette remain unclear, our findings support the need for further investigations into the safety profile of chronic e-cigarette use to better inform regulation and policy.

## Data Availability

The data that support the findings of this study are available from Cedars-Sinai Medical Center, upon reasonable request. The data are not publicly available due to the contents including information that could compromise research participant privacy/consent.

## Acknowledgements

We dedicate this work to memory of Dr. Ronald G. Victor, MD, to whom we are indebted for his pioneering investigations in cardiovascular physiology.

---

**Supplementary Table 1.** Summary of Wilcoxon Signed-Rank Test Between Pre- and Post-Stress Myocardial Blood Measurements Within Groups

| Group | Myocardial Blood | Negative Ranks |  |  | Positive Ranks |  |  | Test Statistics |  |
| --- | --- | --- | --- | --- | --- | --- | --- | --- | --- |
|  |  | *n* | Mean Rank | Sum of Ranks | *n* | Mean Rank | Sum of Ranks | Ties | p-value |
| Controls | Volume (IU) | 5 | 4.8 | 24 | 5 | 6.2 | 31 | 1 | 0.76 <sup>a</sup> |
|  | Flux Rate (s<sup>-1</sup>) | - | - | - | 10 | 5.5 | 55 | 0 | 0.002 * |
|  | Flow (IU/s) | - | - | - | 10 | 5.5 | 55 | 0 | 0.002 * |
| Tobacco users | Volume (IU) | 6 | 5.0 | 30 | 4 | 6.3 | 25 | 0 | 0.846 |
|  | Flux Rate (s<sup>-1</sup>) | 1 | 2.0 | 2 | 9 | 5.9 | 53 | 0 | 0.006 * |
|  | Flow (IU/s) | 2 | 3.0 | 6 | 8 | 6.1 | 49 | 0 | 0.0273 |
| E-Cig users | Volume (IU) | 6 | 5.8 | 35 | 4 | 5.0 | 20 | 0 | 0.492 |
|  | Flux Rate (s<sup>-1</sup>) | 5 | 7.0 | 35 | 5 | 4.0 | 20 | 0 | 0.492 |
|  | Flow (IU/s) | 6 | 6.0 | 36 | 4 | 4.8 | 19 | 0 | 0.432 |

\* Denotes statistically significant changes from pre- to post- test at the alpha = 0.01 level
<sup>a</sup> Based on a normal approximation, due to tie in ranks

